# Inspiring Diverse Researchers in Virginia: Cultivating Research Excellence Through a Career Building Program

**DOI:** 10.1101/2023.11.02.23297785

**Authors:** Lina V. Mata-McMurry, Jennifer V. Phillips, Sandra G. Burks, Adam Greene, Sana Syed, Karen C. Johnston

## Abstract

Historically underrepresented groups in biomedical research have continued to experience low representation despite shifting demographics. Diversity fosters inclusive, higher quality, and innovative team science. One avenue for diversifying research teams is integrating diversity-focused initiatives into Clinical and Translational Science Award (CTSA) Programs, such as the integrated Translational Health Research Institute of Virginia (iTHRIV). In 2020, iTHRIV participated in Building Up, developed by the University of Pittsburgh CTSA, intended to increase representation and improve career support for underrepresented groups in the biomedical workforce. Drawing lessons from this study, iTHRIV implemented the “inspiring Diverse Researchers in Virginia” (iDRIV) program. This year-long program provided education, coaching, mentoring, and sponsorship for underrepresented early-career investigators in the biomedical workforce. To date, 24 participants have participated in the program across three cohorts. Participants have been predominantly female (92%), with 33% identifying as Hispanic/Latinx, 29% as Black, and 13% Asian. Notably, 38% of scholars have subsequently achieved at least one accomplishment, such as receiving a local research honor or award and an extramural funding award from a foundation or federal agency. The iTHRIV iDRIV program serves as a model for providing career support to developing investigators from underrepresented backgrounds, with the overall goal of improving patient health.

## Introduction

The lack of diverse representation in the biomedical research workforce is a critical issue that requires urgent attention and immediate resolution. The National Institutes of Health (NIH) definition of populations underrepresented in biomedical research encompasses individuals from certain racial and ethnic groups, individuals with disabilities, and individuals from socioeconomically disadvantaged backgrounds [1]. Despite the enduring efforts to diversify the field and the shifting U.S. demographics, representation of diverse racial and ethnic groups in the nation’s scientific research faculty positions remains persistently low: only 4% are Black, 5% are Hispanic, 0.2% are Native Americans, and 0.1% are Native Hawaiian, compared to the White majority of 72% [2,3]. Furthermore, the underrepresentation of individuals with disabilities persists, as evident in the persistently low and continually declining number of applicants and recipients with disabilities of NIH-funded grants [4]. Similarly, students from low socioeconomic status backgrounds attain advanced degrees at disproportionately lower rates [5]. Like all underrepresented groups, they often face systemic barriers, limited opportunities for advancement, and implicit biases that hinder their progress in science, technology, engineering, and mathematics (STEM) careers. This underrepresentation perpetuates a cycle of limited role models and mentors for aspiring scientists from diverse backgrounds, further impeding the career journey of underrepresented individuals in biomedical research and contributing to the lack of diverse research teams in STEM [6,7].

Diversity and research excellence are closely intertwined concepts. Diverse research teams are essential, as they bring together unique experiential backgrounds, perspectives, and problem-solving skills [8]. The collective knowledge within diverse teams helps mitigate unconscious biases, enhances creativity, offers a broader range of insights, and generates innovative ideas. Diverse teams lead to higher quality, more rigorous, impactful research, scientific innovation, and discovery [9]. Furthermore, diverse research teams often better address the distinct healthcare needs of diverse patient populations [11,12]. As such, embracing diversity in biomedical research can transform clinical approaches and enhance the research question’s relevance.

The impact of diversifying the biomedical research workforce extends far beyond the research itself; it also directly improves patient care [12,13]. A workforce reflecting the diverse demographics of society enhances the ability to develop interventions and treatment strategies that are more effective and tailored to various populations [14,15]. Fostering the diverse perspectives that drive excellence in the research workforce will generate innovative approaches that will effectively address and eliminate health disparities while providing us with a deeper understanding of the biological, social, and environmental factors contributing to disease outcomes.

Implementing comprehensive and proactive solutions to improve diversity in the field is essential. Solutions include actively recruiting individuals with unique and different perspectives and experiences from underrepresented groups, providing equitable opportunities for training and career development, and fostering inclusive and supportive environments that value diversity [16,17]. Additionally, efforts to dismantle systemic biases and discriminatory practices within the academic and medical sectors are crucial for creating a more inclusive and representative workforce [18].

One avenue for implementing diversity-related initiatives focused on workforce development is to incorporate these into the existing framework of the NIH National Center for Advancing Translational Sciences (NCATS) Clinical and Translation Science Award (CTSA) Program. The CTSA program supports a national network of 62 academic health centers and institutions called “hubs.” These hubs provide resources and support to improve the translational research process, relying on a highly-skilled, creative, and diverse translational science workforce [19]. The integrated Translational Health Research Institute of Virginia (iTHRIV) joined other CTSAs in 2019. It currently includes partners across Virginia, including the University of Virginia (UVA) (Northern and Central VA), Virginia Tech and Carilion Clinic (Southwestern VA), and Inova (Northern VA) [20]. In 2020, iTHRIV joined “Building Up,” a novel program developed by the CTSA hub at the University of Pittsburgh as part of a clinical research study. This program aimed to test a 12-month targeted Career Education and Enhancement for Research Diversity (CEED) intervention designed to build a community of brilliant and underrepresented scholars [21,22]. While the results from the Building Up study have been previously presented (unpublished data) and publication is pending, it demonstrated an effective platform for diversifying the research workforce. The lessons learned from this research were subsequently implemented in iTHRIV’s “inspiring Diverse Researchers in Virginia” (iDRIV) program [23]. This year-long program seeks to jump-start and drive the research journey of early career and aspiring faculty at UVA through education, coaching, mentoring, and sponsorship, designed to support researchers nationally underrepresented in clinical and translational science. Here, we outline our strategy for the ongoing delivery of targeted career development support for promising early-career clinical and translational scientists, emphasizing research excellence through embracing diversity. We describe the first three years of the program.

## Methods

Drawing inspiration from the Building Up study and the CEED program, we tailored and implemented iDRIV at a single academic health system (UVA), closely focusing on our workforce’s specific needs and characteristics. A fundamental pillar of the program was the delivery of guidance from a near-peer mentor per cohort. The near-peer mentor model was first implemented at the Walter Reed Army Institute of Research (WRAIR) for undergraduate research experiences for students in STEM [24]. Near-peer mentors were typically individuals a few years ahead in their career journey and often shared similar backgrounds with their mentees. They experienced both sides of the mentor-mentee relationship, simultaneously advancing their professional paths while offering support, guidance, and relatable advice to their mentees in earlier stages of their careers [24,25]. A program director (SS) and a program manager (JVP) led the program.

### Applicant eligibility criteria, recruiting and application process

The iDRIV program was designed for early-stage investigators committed to pursuing a career in clinical or translational research [26]. Eligibility criteria included candidates who were late pre-faculty (fellows or post-docs) and early career faculty (clinical instructors, assistant professors, or early associate professors [<5 years]). While not used as part of the selection criteria, applications did have candidates self-identify whether they met the NIH definition for underrepresented individuals in biomedical research to better understand current diversity at our institution [1].

Applicants were recruited through various approaches, including direct departmental invitations, email blasts, and postings on the iTHRIV website and the School of Medicine newsletter.

Requests for Applications (RFA) were sent out, and applications were received annually. Applicants were required to submit an application form through the Research Electronic Data Capture tool (REDCap)[27] along with a recent Curriculum Vitae or an NIH biosketch. The application form collected demographic data (some questions were optional), contact information, educational background, current employment information, and specific mentor information. Additionally, we gathered data specific to the applicant’s research–such as their research focus, short and long-term goals, and an outline of prospective challenges that might impact their research–and their expectations for the program. The iTHRIV and iDRIV leadership team subsequently reviewed applications and selected candidates who exhibited the qualities of researchers striving for excellence.

### Program format and sessions outline

Selected iDRIV scholars participated in the yearlong program, with the first three cycles following the 2020-2021, 2021-2022, and 2022-2023 academic years. Each cycle began with individualized meetings between each scholar and the program near-peer mentor to identify and share resources that may have helped support the scholar. 1.5-hour collaborative sessions were held monthly via Zoom. The Building Up cohort started with 12 sessions that included topics such as the importance of mentorship, promoting research on social media, organizational skills, wellness, and work-life balance, among others (**Table 1**). An additional three sessions were developed and added to subsequent cohorts to address the needs and feedback received from past participants. Sessions were a mix of presentations and panel discussions that encouraged interaction between the speakers, near-peer mentor, and scholars. In addition, the monthly collaborative iDRIV sessions were supplemented with an optional 6-week NIH proposal development training series. Each cycle included a networking session that allowed scholars to meet with UVA Health System and University leadership. These networking sessions facilitated an open discussion of the scholars’ research goals and allowed for the exchange of solutions to overcome potential barriers against success.

**Table 1.** Description of Program Sessions.

| **Title** | **Description** |
| --- | --- |
| Orientation and Why Mentoring? | Overview of program, staff, and iTHRIV resources with emphasis on mentorship. |
| Building Your Brand: Promoting Your Research on Social Media | How to increase visibility of a researchers work with social media. Includes a 30-minute panel discussion. |
| Getting Organized | Time management strategies that will increase efficiency as a researcher. |
| Curriculum Vitae (CV), Biosketches, and Grant Submission Process | Review of faculty CV format and the grant submission process at UVA. |
| Contract Negotiating in Academic Medicine | Career success is dependent on negotiation skills that many new researchers do not possess. This session delves the negotiation topic. |
| Scholar Check-in: What challenges are you facing? | Meetings done at the beginning of each program year. |
| Wellness and Work Life Balance | Tips on how to manage stress at work and at home. |
| Networking and Engaging with Leadership | Meeting with Health System and University Leadership to help understand the mission and vision and how the scholars interact with the mission and vision. |
| Giving Effective Presentations | Interactive and didactic session on public speaking and discussing their science. |
| Scholar Presentations | Practice sessions for giving a brief talk. |
| Goal Setting | Writing out goals for next 6 months, 1 year, 5 years and how each scholar will go about achieving them. |
| Library Services and Resources | Resources available to all researchers in the library that are less known but very valuable. |
| Mentoring Matters | The importance of the mentoring relationship and how a team of mentors can be effective. |
| Planning for Promotion or Tenure | Tips for understanding the promotion and tenure process and when to start planning. |
| Microaggressions | Identifying and managing microaggressions in the workplace. |
| Biostatistics Resources | Understanding how to access the biostatistical resources available. |

### Program Evaluation

After completion of the program, an annual optional iDRIV program evaluation survey was distributed to participants. The survey contained a total of 13 questions. The first five questions (**Figure 1**) evaluated participant satisfaction and were presented on a Likert scale (strongly agree, somewhat agree, neither agree nor disagree, somewhat disagree, and strongly agree). Eight open-ended questions inquired about any additional comments, what aspects of the program were most and least beneficial, and suggestions for the program in the future.

**Figure 1.**
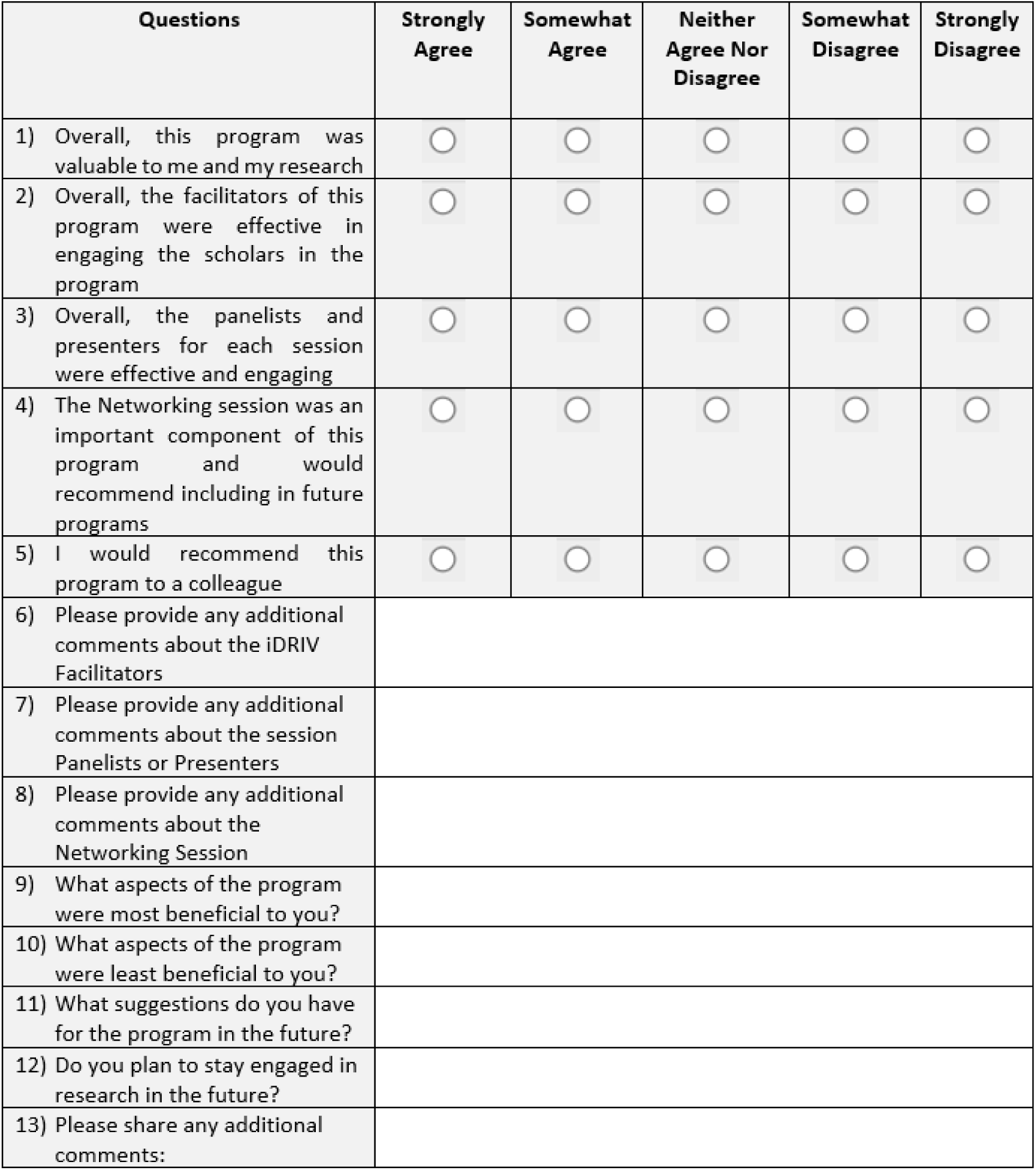
Annual iDRIV Program Evaluation Survey.

As metrics of the scholars’ and overall program success, we gathered information on scholars’ accomplishments and publications for up to three years after their participation. Program data were collected using Qualtrics (Qualtrics, Provo, UT) or REDCap [27]. Descriptive data analysis was prepared for this manuscript.

## Results

Cohort One of the iDRIV program was launched in collaboration with the University of Pittsburgh as part of the Building Up study between October 2020 and July 2021. UVA School of Medicine departments (medicine, pathology, pediatrics, urology, neurology, surgery, and public health sciences) subsequently supported Cohort Two (October 2021 to July 2022) and Cohort Three (October 2022 to July 2023).

### Scholars

A total of 24 scholars were accepted into the three cohorts. Characteristics of iDRIV scholars are presented in **Table 2**. The overall cohort was 92% female, 33% Hispanic/Latinx, 54% White, 29% Black, and 13% Asian. 75% (n=18) were physician-scientists, and 25% (n=6) were Ph.D. researchers. At the time of application, scholars were predominantly fellows/post-docs (54%) and assistant or associate professors (46%). Confirmation of early-stage investigator status was determined not only by the scholar’s institutional position at the time of application but also by the number of years since the highest degree was achieved as defined by the NIH (median=6 years) [26]. A summary of the applicant’s short and long-term goals is listed in **Table 3**. The most common goals at the time of application included publication of manuscripts as first or co-author, presentation at conferences or meetings, successful application for NIH career development awards (K), and successful application for NIH Research Project awards (R01).

**Table 2.** Cohort characteristics of the Inspiring Diverse Researchers in Virginia (iDRIV) Program.

| Characteristic | Total<br>n=24 |  | Cohort<br>One<br>2020-2021<br>n=8 |  | Cohort<br>Two<br>2021-2022<br>n=9 |  | Cohort<br>Three<br>2022-2023<br>n=7 |  |
| --- | --- | --- | --- | --- | --- | --- | --- | --- |
|  | n | % | n | % | n | % | n | % |
| **Gender** |  |  |  |  |  |  |  |  |
| Female | 22 | 92% | 8 | 100% | 8 | 89% | 6 | 86% |
| Male | 2 | 8% | 0 | 0% | 1 | 11% | 1 | 14% |
| **Ethnicity** |  |  |  |  |  |  |  |  |
| Hispanic or Latino/a/x | 8 | 33% | 2 | 25% | 2 | 22% | 4 | 57% |
| Non-Hispanic or Latino/a/x | 16 | 67% | 6 | 75% | 7 | 78% | 3 | 43% |
| **Race** |  |  |  |  |  |  |  |  |
| American Indian or Alaska Native | 0 | 0% | 0 | 0% | 0 | 0% | 0 | 0% |
| Asian | 3 | 13% | 1 | 13% | 2 | 22% | 0 | 0 |
| Black or African American | 7 | 29% | 1 | 13% | 3 | 33% | 3 | 43% |
| Native Hawaiian or Other Pacific Islander | 0 | 0% | 0 | 0% | 0 | 0% | 0 | 0% |
| White | 13 | 54% | 6 | 74% | 4 | 45% | 3 | 43% |
| Prefer not to Answer | 1 | 4% | 0 | 0% | 0 | 0% | 1 | 14% |
| **Degree** |  |  |  |  |  |  |  |  |
| M.D. | 15 | 63% | 5 | 62% | 7 | 78% | 3 | 43% |
| Ph.D. | 6 | 25% | 2 | 25% | 1 | 11% | 3 | 43% |
| M.D./Ph.D. | 3 | 12% | 1 | 13% | 1 | 11% | 1 | 14% |
| **Career Status or Position at Time of Application** |  |  |  |  |  |  |  |  |
| Associate Professor, less than 5 years | 1 | 4% | 0 | 0% | 1 | 11% | 0 | 0% |
| Assistant Professor | 10 | 42% | 1 | 13% | 3 | 33% | 6 | 86% |
| Fellow or Post-doc | 13 | 54% | 7 | 87% | 5 | 56% | 1 | 14% |
| **Department / Institute** |  |  |  |  |  |  |  |  |
| Medicine | 10 | 42% | 5 | 64% | 3 | 34% | 2 | 29% |
| Neurology | 1 | 4% | 1 | 12% | 0 | 0% | 0 | 0% |
| Pathology | 2 | 8% | 0 | 0% | 1 | 11% | 1 | 14% |
| Pediatrics | 5 | 21% | 0 | 0% | 3 | 34% | 2 | 29% |
| Public Health Sciences | 1 | 4% | 1 | 12% | 0 | 0% | 0 | 0% |
| Surgery | 4 | 17% | 1 | 12% | 2 | 21% | 1 | 14% |
| Urology | 1 | 4% | 0 | 0% | 0 | 0% | 1 | 14% |
| Years Since Highest Degree Achieved (median, 25<sup>th</sup>-75<sup>th</sup> percentile) | 6<br>(4-12) |  | 6<br>(3-12) |  | 6<br>(4-8) |  | 10<br>(8-14) |  |

**Table 3.** Summary of scholar’s short and long-term goals at application.

| Summary of scholars short and long-term goals at application* | Responses |
| --- | --- |
| Apply for a career development grants (K, F, T grants)<sup>#</sup> | 20 |
| Present at local, national, international conferences/meetings | 18 |
| Publish manuscripts as first author and co-author | 17 |
| Apply for a Research Project Award (R01) | 9 |
| Career Advancement (Secure a faculty position, advance to associate professor, or get on the tenure track) | 6 |
| Gain and develop grant application knowledge and writing skills | 6 |
| Networking opportunities | 6 |
| Apply for Foundation Grants | 5 |
| Develop a successful and funded research program or lab | 5 |
| Understand the resources that are available at the University to conduct research | 3 |
| Apply for Department of Defense Grant | 2 |
| Complete a Masters | 2 |
| Learn soft skills to promoting oneself, finding our voice, learning to negotiate for better opportunities, and learning to be an effective, yet caring mentor and leader | 2 |
| Apply for a Diversity Supplement Grant | 1 |
| Apply for a Fogarty Fellowship | 1 |
| Apply for a Research Program Project Grant (P01) | 1 |
| Apply for Exploratory/Developmental Research Grant Award (R21) | 1 |
| Apply for internal department grants | 1 |
| Graduate two PhD students in my lab | 1 |
| Work for the World Health Organization conducting research in perinatal health | 1 |

*\*Multiple goals per applicants are indicated*
*<sup>#</sup>Scholars specified goal of applying for K01, KL2, K23, K43, K99/R00 pathway to independence award, Postdoctoral Fellowship (F32), Research Training Grant (T32)*

### Sessions Feedback

The program sessions were well-received overall. Cohorts Two and Three had the opportunity to identify additional session needs, and extra sessions were added to their cohorts. For instance, Cohort Three participants expressed a need for research methods and statistical skills. As a result, a session with resources addressing those needs was provided in that cycle. Cohorts One and Two did not have sessions on Library Services and Resources, Microaggressions, or Biostatistics Resources. Cohort Three did not have sessions on Contract Negotiating in Academic Medicine, Wellness and work-life balance, or Scholar Presentations.

## Evaluation Survey Results

The annual survey was distributed to Cohorts Two and Three (n=16), while the Building Up survey results will be presented separately by the University of Pittsburgh CTSA. In total, 50% (n=8) of iDRIV scholars responded to the annual survey (Cohort Two n=5, Cohort Three n=3).

The respondents to the annual survey (**Figure 2**) overwhelmingly expressed their satisfaction with the program. They strongly agreed that the program was valuable (100%), found the facilitators (88%) and panelists (100%) of each session effective, considered the Networking session an important component of the program (75%), and indicated a willingness to recommend this program to their colleagues (100%).

**Figure 2.**
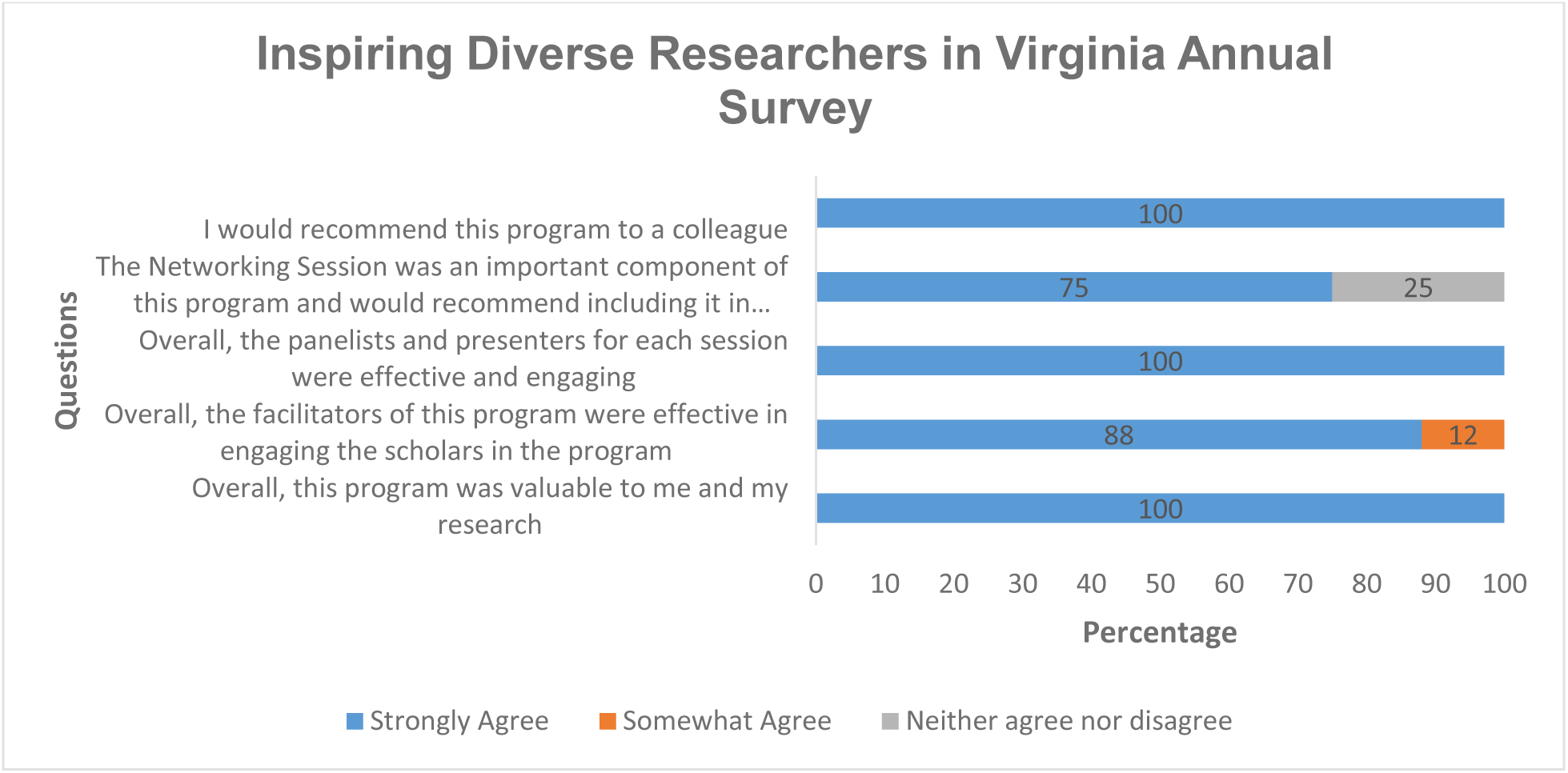
Annual iDRIV Evaluation Survey Questions.

In summary, the open-ended questions revealed that the program was well-rounded, helpful, and provided the necessary resources for a future career in research. The iDRIV facilitators were commended for their outstanding performance, willingness to answer questions, and responsiveness. The panelists were praised for their relatability and unique perspectives on research. The networking sessions, where participants had the opportunity to meet with leadership, were found beneficial. However, one scholar recommended making the session format more informal to encourage open discussions, such as having small tables with food instead of one large conference room table.

Regarding the program’s future, 25% of participants recommended in-person sessions to allow for more interactions among scholars. In response to a question about the most beneficial session of the program, participants commented that all the sessions were beneficial, and the most beneficial sessions were: ‘Orientation and Why Mentoring?’ (25%), ‘Giving Effective Presentations’ (25%), ‘Curriculum Vitae (CV), Biosketches, and Grant Submissions Process’ (13%), ‘Getting Organized’ (13%), and ‘Goal Setting’ (13%). Additionally, 88% of evaluation survey respondents expressed their intention to stay engaged in research in the future.

### Scholars Accomplishments

iDRIV scholars’ accomplishments as of August 2023 are presented in **Table 4**. We considered these as positive outcome measures of the program. In total, 38% (n=9) of all scholars have achieved at least one accomplishment, and one was recognized for receiving three awards. The awards received included the UVA School of Medicine Team Science Award, UVA Department of Internal Medicine Fellow of the Year Award, and membership in the UVA Office of Faculty Affairs and Development Academy of Excellence in Education. These three awards recognized excellence in academic medicine and research.

**Table 4.**
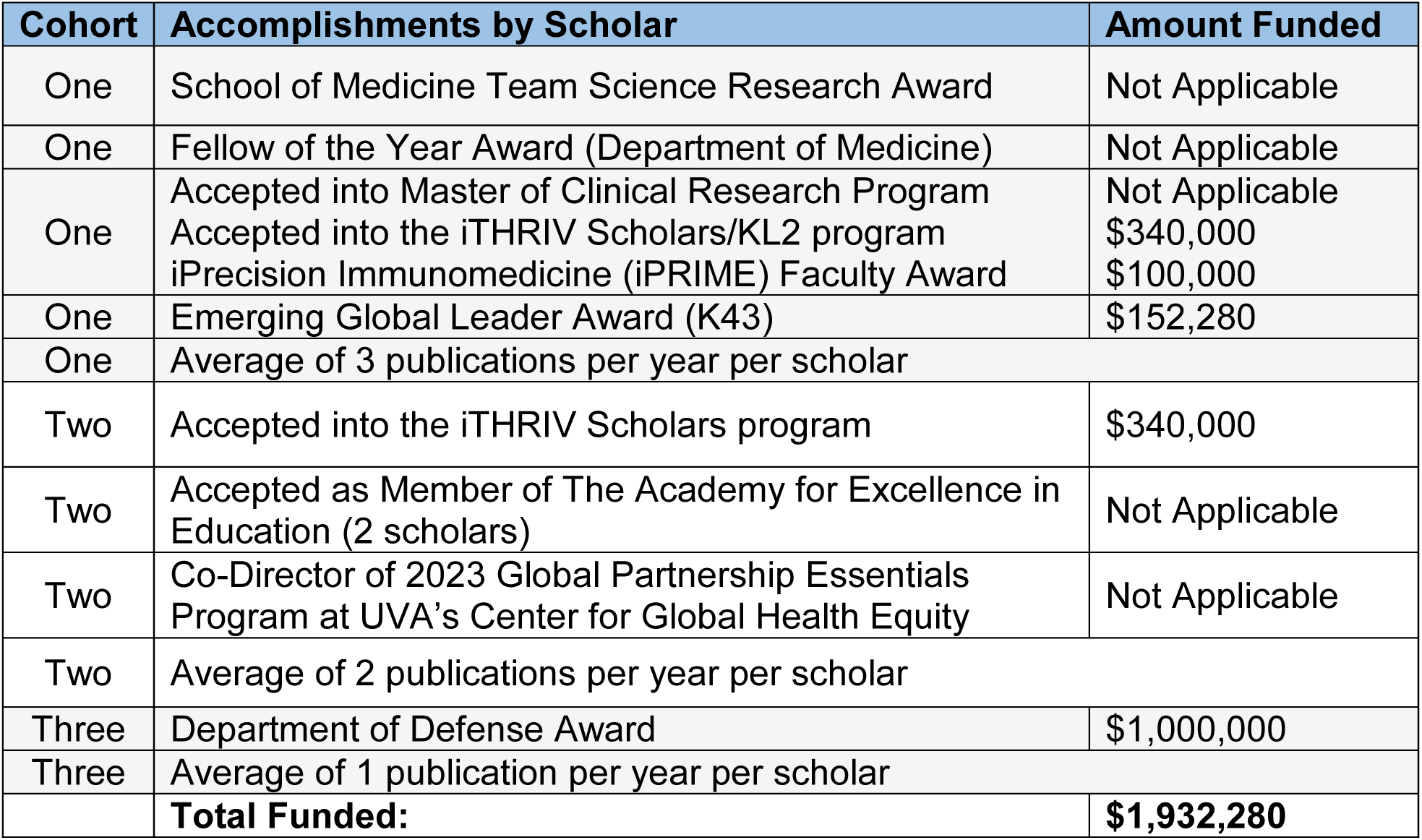
Accomplishments of iDRIV Scholars per Cohort.

Across all three cohorts, four iDRIV scholars have received federal and institutional funding totaling $1,932,280 after participating in iDRIV. Furthermore, we used the average number of publications per cohort scholar per year as another measure of success, with an average publication rate for early-stage investigators considered to be 1-3 papers per scholar per year [28]. Every cohort met this average of 1 to 3 papers published annually (**Table 4**). When evaluating this success metric we did not factor in authorship position or journal quality.

## Discussion

Diverse research teams bring forth new experiences and innovative ideas. However, biomedical research teams lack diversity, perpetuating a cycle of underrepresentation that continues to widen the gap. iTHRIV’s iDRIV program was developed to improve human health by diversifying the biomedical research workforce and providing career support to promising early career scholars who are historically underrepresented in research and translational science. We recognized the urgent need for such a program locally and successfully implemented it at UVA.

Though we do not have a comparison cohort, the success of iDRIV is evident in the achievements and publications of its scholars during and after the program. These achievements align with the key outcomes analysis of the CEED program [22]. The CEED program went a step further by comparing its participants with a control group and found that scholars were more likely to have peer-reviewed publications, and receive career development awards or research project grants. However, these differences did not reach statistical significance.

iDRIV serves as a crucial pathway for our teams to reach the highest level of research excellence, ultimately leading to novel ideas for disease prevention, diagnosis, and treatment. We anticipate that iDRIV will lead to continued successes for our scholars, including but not limited to being provided with continued training opportunities such as the K12 scholars program or receiving an R01 or equivalent grant from the NIH or other governmental institutions. By fostering and graduating new investigators, the program has expanded the pool of diverse future research mentors and leaders, raising a more inclusive and representative biomedical workforce and, generating a more significant impact in our institution and beyond.

## Program Limitations

Limitations of our work include that this is a new program, and our sample is small and lacks long-term outcomes. Additionally, not all of our scholars have provided feedback so our results may be biased by the limited respondents. We do not have a comparison cohort, and selection bias could contribute to our scholar’s success. Since we have accepted all applicants who meet the eligibility criteria, systematic selection bias seems unlikely. It is possible, however, that department leaders have introduced bias by suggesting that only the most successful candidates apply or that most successful scholars are more likely to apply. By continuing to refine our outcome measures and track the successes of our programs, we will improve our ability to accurately assess how participation in iDRIV impacts scholars’ future careers.

One major limitation of this program is the lack of representation in the cohorts from specific groups such as American Indians or Alaska Natives (AIAN) and Native Hawaiians or other Pacific Islanders (NHOPI). As iDRIV directly recruits applicants from UVA, the cohorts roughly reflect UVA demographics. Thus, we are limited in our ability to recruit individuals who are underrepresented not just in NIH programs but who are underrepresented at this institution [29]. Notably, recent efforts have been undertaken by UVA to increase the representation of certain groups, such as American Indians, at the institution as a whole. To this end, UVA has recently established the role of tribal liaison to build relationships with local American Indian groups and encourage members to pursue education at UVA [30]. These initiatives aimed towards diversifying the University demographics will allow us to diversify our cohorts further and thus significantly improve the perspectives and training within our program.

Another limitation pertains to our application process and data collection. Although we request applicants to self-identify as an underrepresented group (yes/no question), we do not collect data on the distribution of disability or socio-economic background of applicants. As a result, we cannot characterize scholars more specifically to determine if this program is effectively reaching individuals with diverse experiences. With this in mind, we are reviewing our process for potential improvement opportunities, such as expanding the self-identification questions. This will allow for a comprehensive view of diversity in our applicants and aid us in recruiting a more representative cohort.

In response to the recent affirmative action ruling by the Supreme Court, our eligibility criteria have been revised [31]. In the next cycle of applications, eligibility will not be assessed based on race in compliance with federal guidelines for college admissions. Instead of asking whether an applicant identifies as underrepresented, future applications will include a new question aimed at capturing candidates’ experiences, encompassing but not limited to their experiences related to being an underrepresented individual and how these experiences have influenced their abilities to contribute to the field.

## Future Directions

As we look to the future, we will continue to review our program design, implementation processes, sustainability plan, and impact to identify areas for improvement. We continue to capture input from our earlier cohorts and incorporate their feedback into plans. Programmatic development is ongoing, and we will establish an external advisory committee to enhance the educational programmatic rigor. We will incorporate in-person sessions to increase participant interaction. Recent additions include a complimentary leadership training program offered to iDRIV scholars. Additional funding sources will be identified to support and expand the program at UVA and extend the program to other iTHRIV partner institutions. Finally, the lessons learned from iDRIV about diversifying the biomedical research workforce will be implemented to address diversity needs across other professional branches of the research workforce, including Clinical Research Professionals and Research Administrators. Our application process will evolve to meet the legal requirements for all iTHRIV programs.

## Conclusion

Establishing career-building programs that support an outstanding group of early-career investigators with varied experiences is imperative for fostering biomedical research excellence. This can directly impact patient care and ensure a more representative and responsive workforce in alignment with our diverse population. The iTHRIV iDRIV program serves as just one model for such a program. Similar programs with novel approaches are warranted to improve diversity in biomedical research and the STEM workforce.

## Data Availability

All data produced in the present study are available upon reasonable request to the authors

## Acknowledgements

We would like to acknowledge the Institute for Clinical Research Education at the University of Pittsburgh Schools of the Health Sciences, along with the dedicated staff of the ‘Building Up’ trial, for serving as the foundation for the development of the iDRIV program. A special note of appreciation to Dr. Doris M. Rubio for her invaluable contributions.

## Ethics Statement

The University of Virginia, Vice President for Research - Institutional Review Board for Health Sciences Research (IRB-HSR) has acknowledged that the work done in the inspiring Diverse Researchers in Virginia (iDRIV) program does not meet the definition of research with human subjects or clinical investigation and therefore is not subject to IRB-HSR oversight.

## Authorship and Contributorship

LM and JP were primary co-authors of the manuscript from the first draft to the final product. KJ, SS, JP, and SB contributed to the conception and adaptation of the iDRIV program. JP managed the program and was responsible for data collection. LM was in charge of data analysis and interpretation. LM, JP, KJ, SS, AG, SB were responsible for critically revising the manuscript. All authors gave their final approval for the version to be published.

